# Microglia transcriptional profiling in major depressive disorder shows inhibition of cortical grey matter microglia

**DOI:** 10.1101/2023.01.11.23284393

**Authors:** Karel W.F. Scheepstra, Mark R. Mizee, Jackelien van Scheppingen, Adelia Adelia, Dennis Wever, Matthew R.J. Mason, Marissa L. Dubbelaar, Cheng-Chih Hsiao, Bart J.L. Eggen, Jörg Hamann, Inge Huitinga

## Abstract

**Background:** Microglia have been implicated in the pathophysiology of major depressive disorder (MDD), but information on biological mechanisms is limited. Therefore, we investigated the gene expression profile of microglial cells in relation to neuronal regulators of microglia activity in well-characterized MDD and control autopsy brains.

**Methods:** Pure, intact microglia were isolated at brain autopsy from occipital cortex grey matter (GM) and corpus callosum white matter (WM) of 13 MDD and 10 age-matched control donors for RNA sequencing. Top differentially expressed genes were validated using immunohistochemistry (IHC) staining. Since gene expression changes were only detected in GM microglia, neuronal regulators of microglia were investigated in cortical tissue and synaptosomes from the cortex by RT-qPCR and Western blot.

**Results:** Transcriptome analysis revealed 92 genes differentially expressed in GM microglia of MDD, but none in WM compared to the control Of these, 81 genes were less abundantly expressed in GM MDD, including C*D163, MKI67, SPP1, CD14, FCGR1A/C*, and *C1QA/B/C*. Accordingly, pathways related to effector mechanisms, such as the complement system and phagocytosis were differentially regulated in GM microglia in MDD. IHC staining revealed significantly lower expression of CD163 protein in MDD. Whole tissue analysis showed an increase in *CD200* (p<0.001) and *CD47* (p=0.068) mRNA, and CD47 protein was significantly elevated (p<0.05) in synaptic fractions of MDD cases.

**Conclusions:** Transcriptional profiling indicates an immune-suppressed microglial phenotype in MDD, possibly caused by neuronal regulation.

## Introduction

Major depressive disorder (MDD) is a significant contributor to the global burden of disease and a leading cause of disability worldwide(1). Insight into disease pathophysiology and novel therapeutics are urgently needed, as treatment resistance is common and occurs in up to 30% of MDD patients(2). Among others, inflammation is a prominent hypothesis in the neurobiology of depression, based on altered levels of pro- and anti-inflammatory cytokines and chemokines in blood and post-mortem brain tissue in MDD(3–5). Furthermore, a higher prevalence of MDD has been associated with chronic inflammatory diseases, such as rheumatoid arthritis, inflammatory bowel diseases, and multiple sclerosis(6,7). Peripheral and central inflammation alters microglia activity(8–10), which may alter neuronal functioning through mechanisms such as synapse stripping(11). Indeed, recent studies show an altered immune status of microglia in MDD(12,13).

Microglia account for 10 to 15% of the cells in the brain and influence major aspects of neuronal functioning. By surveilling the brain for debris, excessive proteins, dysfunctional synapses, and aberrant neurons, they regulate synaptic transmission and neural plasticity in health and disease(14). In psychiatric disorders, microglial changes have been demonstrated in schizophrenia, autism spectrum disorder, MDD, and bipolar disorder(13,15,16). However, it remains uncertain whether – and in which state – microglia have beneficial or detrimental effects in different neuropathological conditions. Studies in MDD report contradicting results, ranging from reactive microglia to microglia in a homeostatic state. Torres-Platas et al. showed vascular infiltration of macrophages and reactive/primed microglia in the white matter of the dorsal anterior cingulate cortex (dACC) of brains of donors with MDD that committed suicide(17). In contrast, a positron emission tomography (PET) study with a radio-labeled tracer for translocator protein (TSPO) – a marker for microglial activation – showed no increase of reactive microglia and even lower TSPO levels in mild to moderate MDD(18). Another TSPO study showed increased binding in the dACC, exclusively in moderate to severe MDD patients with suicidal thoughts(19). Finally, two recent post-mortem studies showed a non-inflammatory signature of microglia, with upregulated homeostatic molecules (TMEM119, CX3CR1, and CD195) and downregulated immune activation markers (CD163, CD14, CD68, and HLA-DR)(12,13).

These contradictory findings regarding microglia in MDD prompted us to investigate their MDD-associated properties. Here, we investigated the transcriptome of primary human microglia in the tissue of a clinically well-characterized MDD brain donor cohort and gene and protein expression of neuronal regulators of microglia in an independent cohort.

## Methods and Materials

### Human post-mortem tissue

Human brain occipital cortex grey matter (GM) and corpus callosum white matter (WM) was provided by the Netherlands Brain Bank (Amsterdam, The Netherlands; https://www.brainbank.nl). Informed consent to perform the autopsy and use tissue and clinical data for research purposes were obtained from all donors,.All NBB procedures were approved by the Medical Ethics Committee of the Amsterdam UMC (locationVUmc, Amsterdam, The Netherlands). For RNA sequencing, GM and WM tissue blocks from the occipital cortex and corpus callosum of MDD donors (n=13) and age-matched non-neurological control donors (n=10) were collected. Non-neurological control donors with cognitive problems, based on clinical data, were excluded from the analysis. Donor characteristics are summarized in **Table 1** and displayed in detail in **Supplementary Table 1**.

**Table 1.** Donor characteristics for MDD and control donors used for RNA sequencing of isolated microglia.

| Diagnosis | n | Sex | Age | PMD | pH CSF |
| --- | --- | --- | --- | --- | --- |
| Control | 10 | 5 m/5 f | 75.0 ± 3.6 | 7.2 ± 0.5 | 6.8 ± 0.1 |
| MDD | 13 | 6 m/7 f | 63.7 ± 4.0 | 8.0 ± 0.7 | 6.7 ± 0.1 |

Data are represented as mean with SEM. Age: age at death (years), PMD: post-mortem delay (hours), CSF: cerebrospinal fluid, m; male, f: female.

For immunohistochemistry (IHC) and quantitative RT-PCR, a second and third cohort with GM tissue blocks from the occipital cortex of MDD donors and age-matched non-neurological control donors was explored. Detailed donor characteristics are displayed in **Supplementary Table 2**.

### Microglia isolation

Corpus callosum and occipital cortex tissue blocks of 5-15 g were dissected at autopsy and stored in Hibernate A medium (Invitrogen, Carlsbad, CA, USA) at 4°C until further processing. Microglia isolations were performed as described before(20,21). Briefly, brain tissue was mechanically dissociated using a tissue homogenizer (VWR, Radnor, PA, USA), followed by enzymatic digestion with trypsin for 45 min at 37°C in Hibernate A medium supplemented with DNAse I (Roche, Basel, Switzerland). Percoll (GE Healthcare, Little Chalfont, UK) density centrifugation was performed, and glial cells were collected from the interlayer. Magnetic-activated cell sorting was performed for positive selection of microglia using anti-CD11b beads (Miltenyi Biotec, Bergisch Gladbach, Germany). Viable cells were counted and the cell pellet was stored in 1 ml cold TRIsure (Bioline, London, UK) at -80°C for further analysis. To assess the purity of isolated cells, CD45, CD11b, and CD15 expression were analyzed by flow cytometry (**Supplementary Figure 1**). In line with previous reports, WM and GM microglia expressed comparable levels of CD11b, while CD45 expression was higher in WM than in GM microglia (20,21).

### RNA isolation

RNA isolation was carried out by QIAzol Lysis Reagent (Qiagen, Hilden, Germany) according to the manufacturer’s protocol using phase separation by addition of chloroform and centrifugation, followed by overnight precipitation in isopropanol at -20°C. RNA concentration was measured using a Nanodrop (ND-1000; NanoDrop Technologies, Rockland, DE, USA), and RNA integrity was assessed using a Bioanalyzer (2100; Agilent Technologies, Palo Alto, CA, USA).

### RNA sequencing

The NEBNext Ultra Directional RNA Library Prep Kit from Illumina (San Diego, CA, USA) was used to process the samples (GenomeScan, Leiden, The Netherlands). Sample preparation was performed according to the protocol of NEBNext Ultra Directional RNA Library Prep Kit from Illumina (NEB #E7420). Briefly, rRNA was depleted from total RNA using the rRNA depletion kit (NEB# E6310). After fragmentation of the rRNA-reduced RNA, cDNA synthesis was performed. This was used for ligation with the sequencing adapters and PCR amplification of the resulting product. The quality and yield after sample preparation was measured with the Fragment Analyzer (Agilent Technologies). The size of the resulting products was consistent with the expected size distribution (a broad peak between 300-500 bp). Clustering and DNA sequencing using the Illumina NextSeq 500 was performed according to manufacturer’s protocols. A concentration of 1.6 pM of DNA was used. NextSeq control software 2.0.2 was used. Image analysis, base calling, and a quality check were performed with the Illumina data analysis pipeline RTA v2.4.11 and Bcl2fastq v2.17. Reads were aligned using the human assembly GRCh37.75. The reads were mapped to the reference sequence using a short read aligner based on Burrows—Wheeler Transform (Tophat v2.0.14) with default settings.

### RNAseq data analysis

Sequencing data were normalized (FPKM), and differential expression was assessed using DESeq2 (https://bioconductor.org/packages/release/bioc/html/DESeq2.html). The read counts were loaded into the DESeq2 package v1.14.1 within the R platform v3.3.0 to find differentially expressed (DE) genes between pre-defined sample groups. The output of this analysis was used to feed all three separate gene regulatory pathway enrichment tools. Post-mortem factors (age, PMD, and pH) were plotted against the top 10 DE, which did not show any correlations.

Ingenuity pathway analyses were performed using the Qiagen IPA tool (Qiagen) and ErmineJ software (https://erminej.msl.ubc.ca/), using all DE genes with an adjusted p-value <0.05 to ensure the qualitative network analysis. Weighted gene correlation networks were identified by weighted gene co-expression network analysis (WGCNA) (version 1.64) library in R (version 3.5.1). Data was converted into a matrix (DESeq2, version 1.20) where transcripts with a sum lower than 10 counts (all GM samples) were considered lowly expressed and removed from the data frame. The variability among the samples was reduced by batch correction using sva (version 3.28.0)(22), where the number of the surrogate variables was predicted using the ‘leek method’(23). Genes with a mean count per million (CPM) expression value in the lower-quartile range (25%) were removed to generate a signed network (beta value of 8) which was then used to perform hierarchical clustering on a topological overlap matrix. Only modules with 100 genes or more were merged with the mergeCloseModules (cutHeight = 0.25) function in the WGCNA package. The module eigengenes were recalculated to their corresponding modules and then used to calculate the module–trait correlation between modules and sample traits (age, gender, and cohort). In the end, the userListEnrichment function was applied, which provides cell type-specific gene expression profiles to compare the identified modules to previously observed networks. In addition, Metascape was used to annotate gene ontology terms for each of the identified WGCNA modules(24).

### Immunohistochemistry

8-µm formalin-fixed, paraffin-embedded (FFPE) sections were cut, and antigen retrieval was accomplished in citrate buffer pH 6.0 (microwave, 10 min at 700 W). Endogenous peroxidase activity was blocked with 1% H_2_O_2_ for 20 min and aspecific binding of secondary antibodies with 10% normal horse serum for 30 min. Sections were incubated with primary antibody (EDHu-1, 1:200; Novus Biologicals, Centennial, CO, USA) overnight at 4°C. The appropriate biotinylated (horse anti-mouse) secondary antibody was applied, followed by conjugation with avidin–biotin horseradish peroxidase complex (Vector Elite ABC kit; Vector Laboratories, Newark, CA, USA). Visualization was established with 3,30-diaminobenzidine chromogen (DAB), and samples were counterstained with hematoxylin.

Images of stained tissue sections were taken using an Axioscope microscope Z1 (Zeiss, Oberkochen, Germany) while using a Plan-Apochromat 20x/0.8 objective with a bright field camera (HV-F203SCL; Hitachi, Tokyo, Japan). Layer I until VI of the cortex and WM were manually outlined with Qupath 0.3.2 software. The DAB background (color deconvolved) was measured for all sections. Subsequently, the DAB threshold for a positive signal was set at four times the average background. DAB particles with size between 10 and 100 mm^2^, exceeding the DAB threshold, were counted as CD163^+^ cells .

### Quantitative RT-PCR

cDNA synthesis was performed using the Quantitect reverse transcription kit (Qiagen) according to manufacturer’s instructions, with a minimal input of 200 ng total RNA. Quantitative PCR (qPCR) was performed using the 7300 Real Time PCR system (Applied Biosystems, Foster City, CA, USA) using the equivalent cDNA amount of 1–2 ng total RNA used in cDNA synthesis. SYBRgreen mastermix (Applied Biosystems) and a 2 pmol/ml mix of forward and reverse primer sequences were used for 40 cycles of target gene amplification. For primer sequences, see **Supplementary Table 3**. Expression of target genes was normalized to the average cycle threshold of *GAPDH* and *EF1A*. Cycle threshold values were assessed with SDS software (Applied Biosystems).

### Synaptosome isolation

Synaptosome fractions were isolated from frozen cortex tissue according to a public protocol from the Kelsey Martin lab that was adapted from Carlin et al.(25). In short, human cryostat sections (50 µm) were collected and stored in a disposable petri dish on dry ice. Sections were homogenized using a dounce homogenizer in a sucrose-rich buffer with protease inhibition. Homogenates were spun down, pelleted, washed, and spun down at 1,400 g for 10 min to remove nuclei. Upon pelleting mitochondria and synaptosomes at 13,800 g for 10’, the pellet was resuspended in sucrose-rich buffer (B) and layered on a gradient of 1.2 M, 1.0 M, 0.85 M sucrose buffer. After ultracentrifuge spinning at 82,500 for 2 h, the top layer between 1.0 M and 1.2 M was aspirated off and processed further for protein detection.

### Western blotting

Isolated synaptosomes or brain tissue cryosections were lysed in cold RIPA buffer containing 50 mM Tris-HCl, pH 8.0, 150 mM NaCl, 1% Nonidet-P40, 0.5% sodium deoxycholate, 0.1% sodium dodecyl sulfate, and protease inhibitors (cOmplete EDTA-free; Sigma-Aldrich, Saint Louis, MI, USA). Protein content was determined using the Pierce BCA Protein Assay kit (Thermo Fisher Scientific, Waltham, MA, USA). For electrophoresis, proteins were separated by sodium dodecyl sulfate-polyacrylamide gel electrophoresis (SDS-PAGE) and transferred to nitrocellulose membranes (Millipore, Bedford, MA, USA) by electroblotting for 90 min (Transblot SD; BioRad, Hercules, CA, USA). Blotted membranes were incubated in blocking buffer (5% BSA in TBS with 0.05% Tween-20 (TBS-T)) for 1 h, followed by o/n incubation at 4°C with primary antibodies directed against CD47 (B6H12.2; Novus Biologicals), CD200 (AF2724; R&D Systems, Minneapolis, MN, USA), PSD95 (810401; Biolegend, San Diego, CA, USA), or β-actin (AC-74; Sigma-Aldrich) in blocking buffer. After several washes with TBS-T, membranes were incubated with DyLightℲ 649 (BioLegend) or IR800 (LI-COR, Lincoln, NE, USA)-conjugated secondary antibody for 1 h at RT (1:2,000 in blocking buffer). After washes in TBS-T, fluorescent signal was detected using the LI-COR Odyssey 9120 Imaging System (LI-COR).

### Statistical analysis

Statistical analysis of RT-qPCR, Western blot and IHC experiments was performed using GraphPad Prism version 9 software (GraphPad Software Inc, La Jolla, CA, USA). Results are shown as mean with standard error of the mean. Statistical analysis was performed using either parametric or non-parametric testing based on the outcome of the Shapiro–Wilk normality test. The applied test for each calculated value is described in the figure legends. P values <0.05 were considered to indicate a significant difference.

## Results

### Microglia isolation yields pure populations of intact microglia

In this study, we isolated intact microglia using a rapid isolation protocol from human occipital cortex (GM) and corpus callosum (WM) (**Figure 1A**). Flow-cytometric phenotyping of intact microglia indicated no differences in CD11b expression, viability, and purity of microglia between MDD (n=13) and control (n=10) donors (**Supplementary Figure 1)**. Membrane CD45 detection was lower in GM microglia isolated from MDD donors (p=0.0141), and for WM microglia, a trend towards decreased membrane CD45 in MDD was visible (p=0.0596) (**Figure 1B**). RNA sequencing of isolated microglia from control GM or WM revealed enrichment of established microglial marker genes (*ADGRG1, CSF1R, P2RY12, C1QA, CX3CR1*) and no notable detection of neuronal (*MAP2, STMN2, SYN1, VGLUT1*), astrocytic (*AQP4, GFAP, ALDH1L1*), oligodendrocytic (*PLP, SOX10, MOG*), or endothelial cell (*CLDN5*) marker genes (**Figure 1C)**. As expected(20,26), hierarchical clustering of gene expression data showed that control GM and WM microglia are phenotypically distinct (**Supplementary Figure 2**).

**Figure 1.**
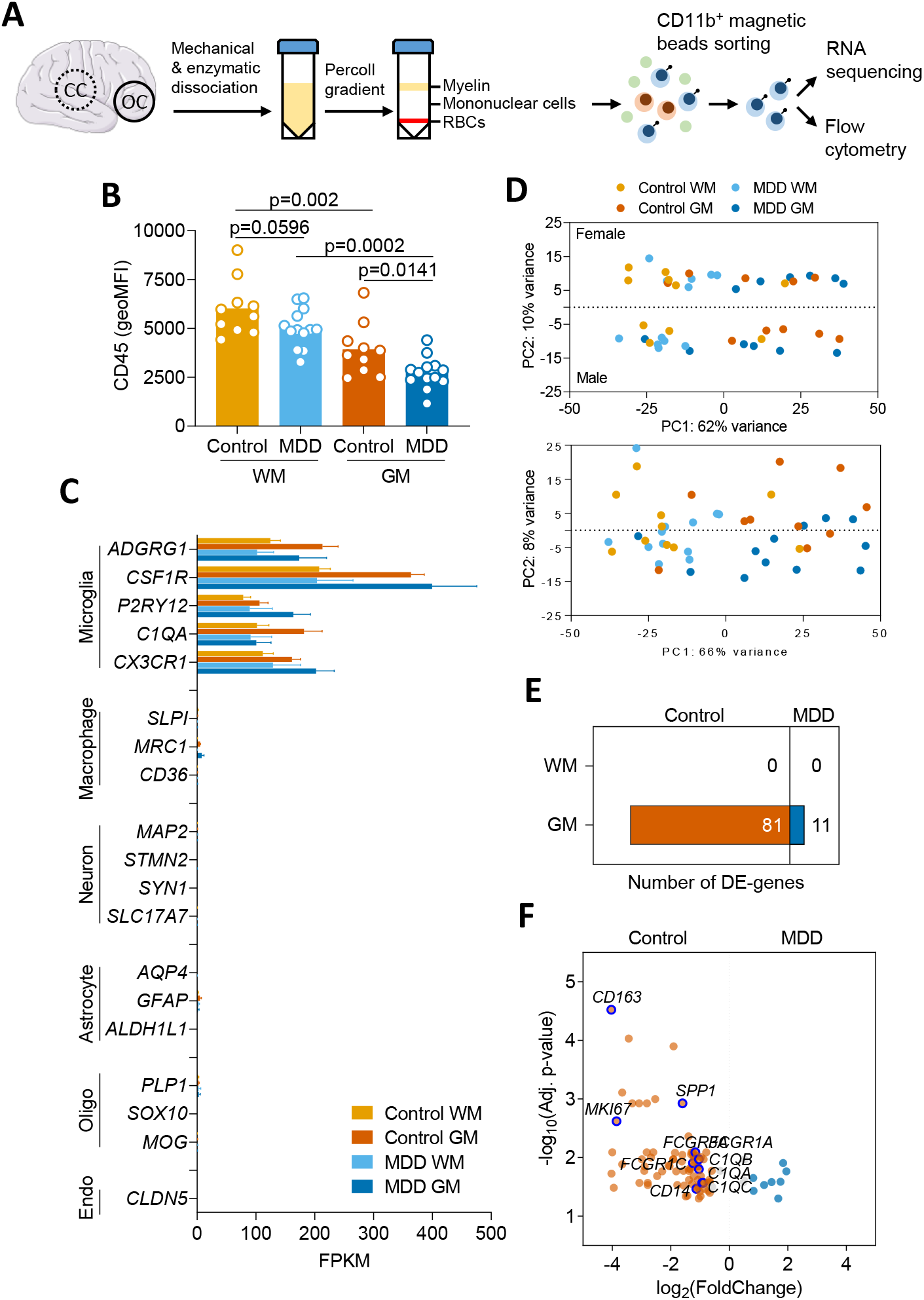
Differential gene expression in microglia isolated from MDD and control brain tissue determined by RNA sequencing. (A) Acutemicroglia isolation from white matter (corpus callosum; CC) and grey matter (occipital cortex; OC) of 13 MDD and 10 age-matched control donors by sorting CD11b^+^ magnetic beads. Flow-cytometric analysis was performed, and RNA was isolated for RNA sequencing. (B) CD45 expression on GM and WM microglia, detected by flow cytometry directly after isolation. Data passed the Kolmogorov-Smirnov normality test, and differences between MDD and control microglia were assessed using unpaired t-test. (C) Normalized expression of genes associated with microglia, neurons, astrocytes, oligodendrocytes, or endothelial cells in control WM or GM microglia. (D) PCA of gene expression demonstrates apparent differences between microglia from GM and WM, and minor differences between microglia derived from male or female donors. (E) Number of DE genes for MDD vs. control microglia for both WM and GM. (F) Volcano plot of DE genes between microglia obtained from MDD or control donors. Threshold indicates DE genes with fold change >1.5 and purple circles the notable downregulated genes of particular interest. DE was determined using an adjusted p-value (false discovery rate) <0.05. RBCs, red blood cells.

**Figure 2.**
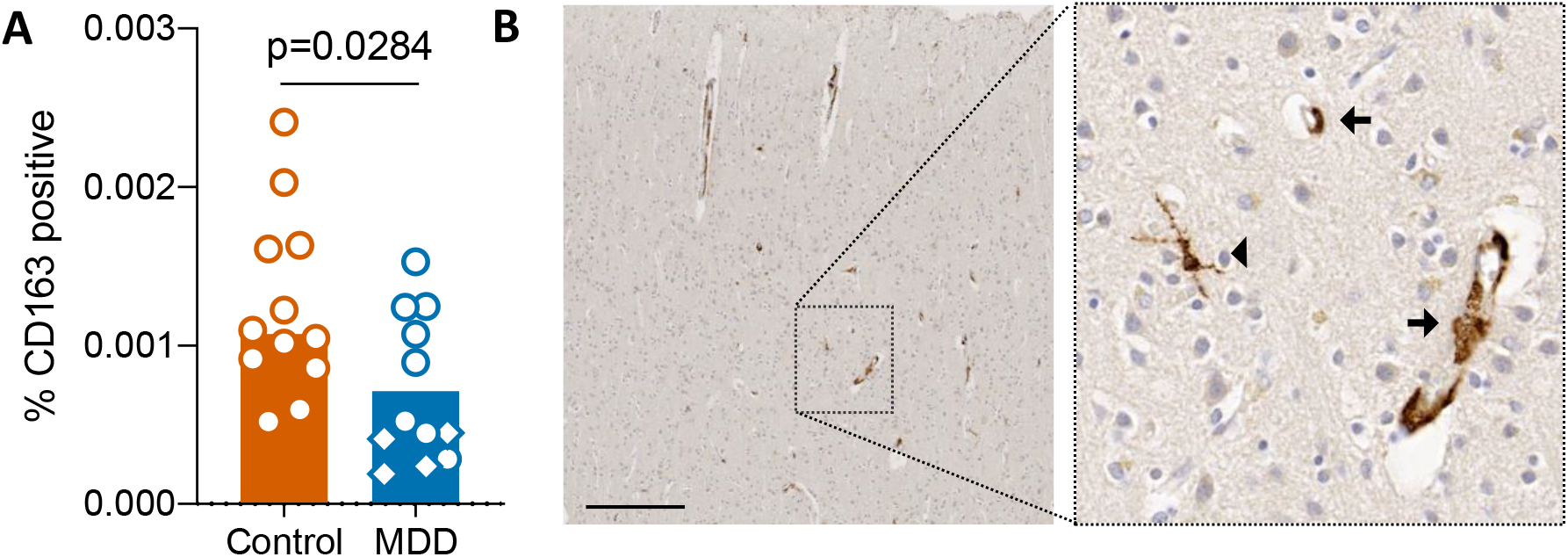
Immunohistochemical staining of CD163. An independent validation cohort of 12 MDD and 12 age-matched control donors was selected, and FFPE tissue was extracted from the occipital cortex GM. (A) Immunoreactivity of CD163, presented as relative expression to total surface. Diamond-shaped symbols indicate donors with a definite depressed state when deceased. (B) Representative images of CD163 staining of a control donor (40× magnification, scale bar = 250 μm), showing CD163^+^ microglia (arrowhead) and perivascular macrophages (arrow). (C/D) Correlation plots showing age, Braak, and amyloid stages in relation to immunoreactivity of CD163. Mann-Whitney U test: *p<0.05.

### Microglial samples cluster based on region and sex

Principal component analysis (PCA) of microglial gene expression data of all samples revealed segregation of the samples in four groups, showing that the largest part of the variation is explained by regional differences between GM and WM microglia (approximately 62%, Figure 1D). Interestingly, the variance in PC2 is defined by sex (10%), with a similar effect in both GM and WM microglia. Further examination of DE genes between males and females only revealed Y chromosome genes, and the X chromosome genes XIST and TSIX, for which expression from the inactive X chromosome accounts for the differential expression. Hence, no further analysis of these DE genes was performed. Cluster analysis of control samples only showed strong clustering of GM and WM samples; however, no complete segregation was seen. No apparent clustering of MDD or control samples was found in either GM or WM.

### GM microglia show differential gene expression in MDD donors

When comparing control GM and WM microglia using DESeq2 analysis, 2,373 DE genes were found, as expected based on previous findings(26), of which 1,691 were higher expressed in WM microglia- and 682 were higher expressed in GM microglia (data not shown). When comparing GM microglia isolated from MDD brains with control GM microglia, 92 DE genes passed multiple testing correction and fold change cutoff, of which 81 were lower and 11 were higher expressed in MDD compared to control microglia (**Figure 1E/F)**. Notable down-regulated genes encoded the scavenger receptor CD163, the proliferation marker Ki-67, the extracellular matrix protein osteopontin, the lipopolysaccharide co-receptor CD14, the high-affinity immunoglobulin gamma Fc receptor (CD64), and complement component C1q chains (see **Supplementary Table 4** for a complete list of all DE genes)(27,28). Regarding microglia isolated from WM, no DE genes passed multiple testing corrections between MDD and control donors.

To validate the top DE gene on protein level, CD163 IHC staining was performed in occipital cortex (GM) of an independent, age-matched cohort of MDD (n=12) and control (n=12) donors. CD163 was expressed by microglia and perivascular macrophages at lower levels in MDD (**Figure 2A/B**, p=0.028). Interestingly, CD163 staining intensity was lowest in donors with a certain depressed state at decease (**Figure 2A**, diamonds).

### Effector function pathways are suppressed in GM MDD microglia

Ingenuity pathway analysis (IPA) was used to identify the genetic regulatory pathways associated with the DE genes in MDD compared to control GM microglia (**Table 2** and **Figure 3**). The largest part of implicated pathways showed lower activity in MDD GM microglia, with the top pathways being the ‘complement system’ and ‘Fc gamma receptor-mediated phagocytosis’. Overlap in regulated genes between overrepresented pathways pointed to suppressed effector functions, specifically phagocytic activity of microglia, as indicated by ‘nitric oxide (NO) and reactive oxygen species (ROS) production by macrophages’, ‘pattern-recognition of bacteria and viruses’ (indicating complement functioning), ‘IL-12 signaling and production in macrophages’, ‘phagosome formation’, and ‘ceramide degradation’ (indicating cell membrane remodeling).

**Table 2.** Ingenuity pathway analysis (IPA) reveals differentially expressed signaling pathways between GM microglia from MDD and control donors.

| Ingenuity canonical pathways | $-\log_{10}$ (adjusted p-value) | Ratio | Genes |
| --- | --- | --- | --- |
| Complement system | 3.43 | 0.0811 | *C1QC, C1QA, C1QB* |
| Fcy receptor-mediated phagocytosis in macrophages and monocytes | 3.38 | 0.0435 | *NCK2, ARPC2, FCGR1A, FCGR3A/FCGR3B* |
| Production of nitric oxide and reactive oxygen species in macrophages | 3.06 | 0.0258 | *TLR2, MAP3K6, MAP3K8, SERPINA1, JAK3* |
| Role of pattern recognition receptors in recognition of Bacteria and viruses | 2.73 | 0.0292 | *TLR2, C1QC, C1QA, C1QB* |
| Pyridoxal 5'-phosphate salvage pathway | 2.71 | 0.0462 | *MAP3K6, PIM1, MAP3K8* |
| Salvage pathways of pyrimidine ribonucleotides | 2.22 | 0.0309 | *MAP3K6, PIM1, MAP3K8* |
| iNOS signaling | 1.9 | 0.0444 | *CD14, JAK3* |
| Phagosome formation | 1.85 | 0.0227 | *TLR2, FCGR1A, FCGR3A/FCGR3B* |
| Iron homeostasis signaling pathway | 1.81 | 0.0219 | *JAK3, CD163, SLC11A1* |
| CD27 signaling in lymphocytes | 1.76 | 0.0377 | *MAP3K6, MAP3K8* |
| IL-12 signaling and production in macrophages | 1.74 | 0.0205 | *TLR2, MAP3K8, SERPINA1* |
| Actin nucleation by ARP-WASP complex | 1.63 | 0.0323 | *NCK2, ARPC2* |
| Ceramide biosynthesis | 1.58 | 0.143 | *SPTLC2* |
| Ceramide degradation | 1.58 | 0.143 | *ACER3* |
| TREM1 signaling | 1.48 | 0.0267 | *TLR2, NLRC5* |
| Sphingosine and sphingosine-1-phosphate metabolism | 1.47 | 0.111 | *ACER3* |
| Toll-like receptor signaling | 1.47 | 0.0263 | *TLR2, CD14* |
| VDR/RXR activation | 1.45 | 0.0256 | *SPP1, CD14* |
| Calcium transport I | 1.43 | 0.1 | *ATP2B4* |
| Dendritic cell maturation | 1.42 | 0.0154 | *TLR2, FCGR1A, FCGR3A/FCGR3B* |

The ratio presented is defined as the number of the differentially expressed genes over the total number of genes involved in each of the pathways.

**Figure 3.**
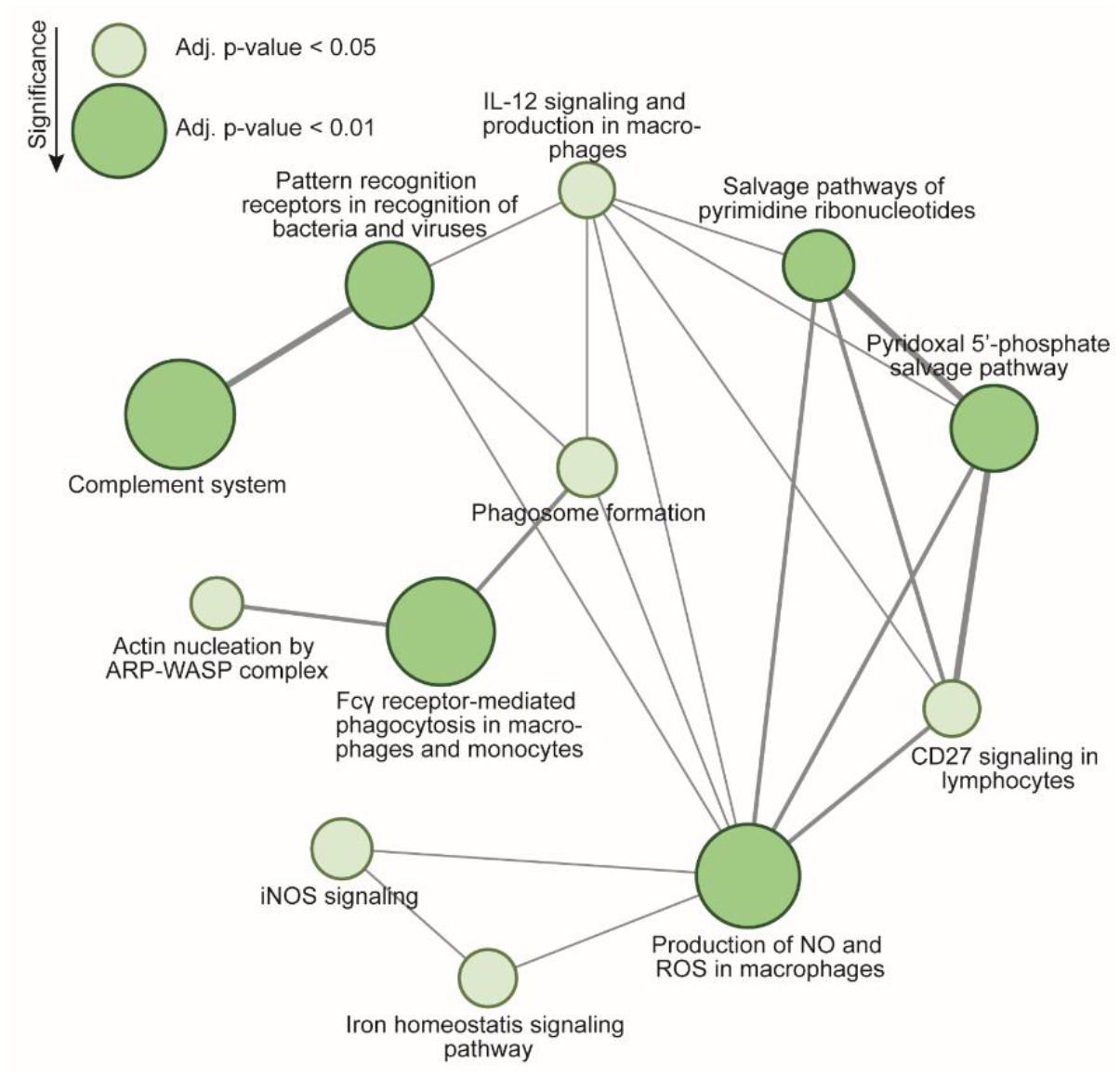
Map of enriched signaling pathways derived from the DE-gene list between GM microglia from MDD and control donors, as determined by ingenuity pathway analysis (IPA). Increasing node size indicates lower adjusted p-value. Light green indicates an adjusted p-value <0.05; dark green indicates an adjusted p-value <0.01. Thicker lines between the nodes indicate a higher number of common DE genes between two pathways.

The gene score resampling (GSR) approach gave non-directional identified pathways that match overrepresented pathways listed in the IPA analysis. GO-terms include ‘IgG binding’, ‘macrophage activation’, ‘Fc-gamma receptor signaling pathway’, ‘regulation of phagocytosis’, ‘plasma membrane invagination’, ‘phagocytosis, engulfment’, and ‘regulation of complement activation’ **(Supplementary Table 5)**. The comparison between WM microglia derived from either MDD or control that yielded no DE genes through DESeq2 analysis similarly only yields one significant hit in the ErmineJ analysis (with false discovery rate <0.05): ‘extracellular matrix binding’.

WGCNA identified clusters of co-expressed genes and allowed for multi-factorial association analysis. When applied to the GM microglia samples with a minimum cluster size of 100 genes, WGCNA revealed 18 distinct modules (after merging related modules), which were further investigated in their association with three different group parameters: age, sex, and cohort (control or MDD) **(Figure 4A)**. Module–trait relationship analysis showed 6 modules significantly related to cohort, 4 modules particularly related to age, and 2 modules especially related to sex. Of the cohort-related modules, 3 modules (M2, M4, and M9) were unique and not associated with age or sex, showing MDD-specific effects on overall gene expression patterns. Module 2, having the highest significant difference relating to cohort, was of particular interest and included microglia function-related GO terms, such as ‘vesicle organization’ and ‘cytosolic transport’ (**Figure 4B)**.

**Figure 4.**
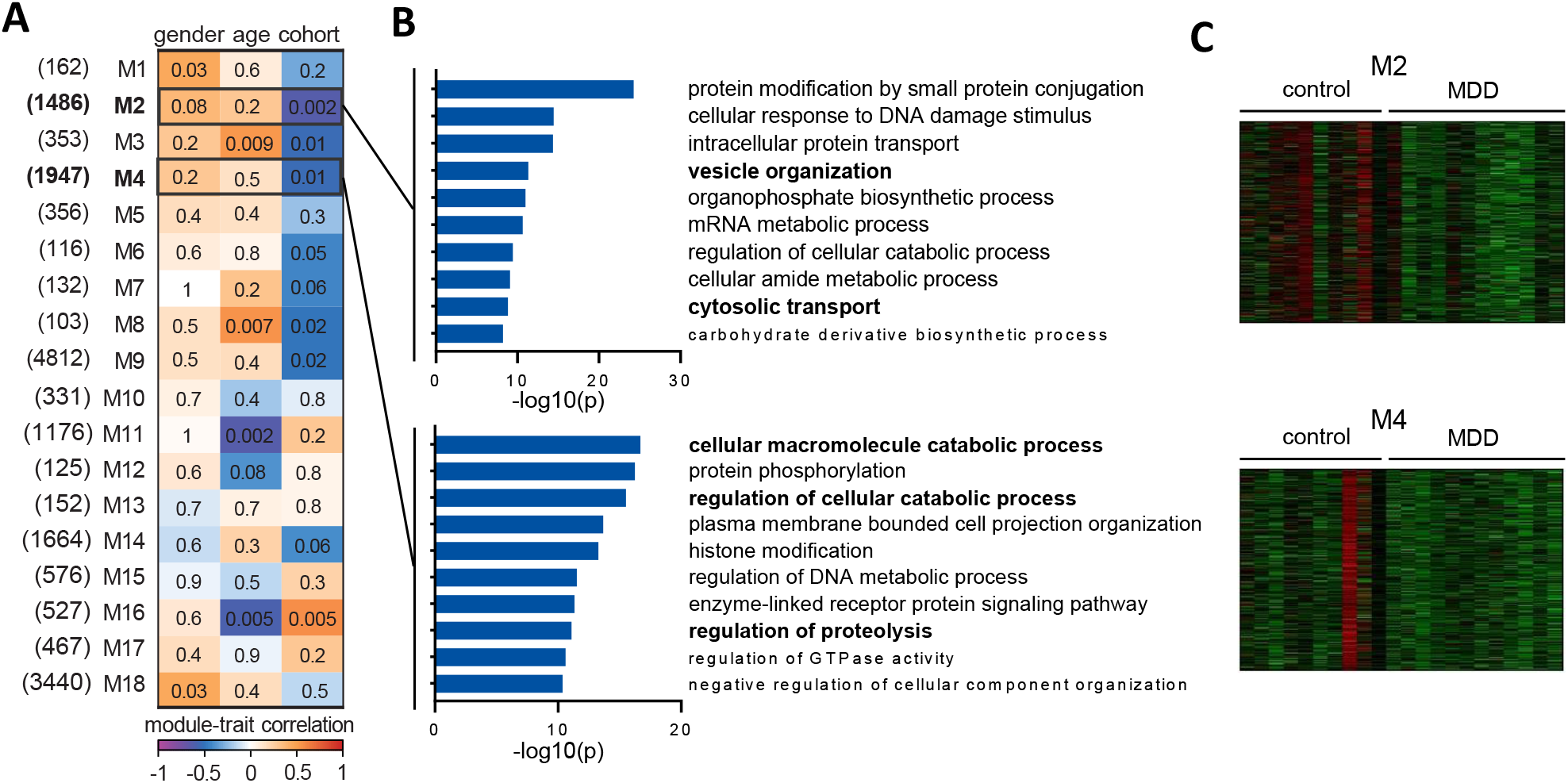
Co-expression networks for GM microglia from MDD or control donors were determined by weighted gene correlation network analysis (WGCNA). **(A)** Eighteen modules identified are provided with their module–trait relationships and the number of genes that belong to each module displayed between brackets. Numbers in the heat map indicate the p-value for the module-trait correlation. (B) A highlight of GO terms (metascape.org) and (C) heatmaps of modules M2 and M4, corresponding to overrepresented pathways in IPA and ErmineJ analyses. Minimum cluster size was set at 100 genes.

### Increased suppression of microglia in MDD GM

To explain a lower activation status of GM microglia in MDD, we examined the expression levels of CD47 and CD200, two well-known proteins involved in neuron-to-microglia signaling that silence microglial immune responses and prevent synaptic pruning(29–31). An independent cohort selection of MDD (n=13) and age-matched control (n=13) occipital cortex (GM) samples were used to investigate *CD47* and *CD200* expression by RT-qPCR **(Figure 5A**). While *CD47* showed a trend towards increased expression in MDD (p=0.068), *CD200* was significantly higher expressed in MDD GM compared to control (p=0.0009) (**Figure 5B**). Since phagocytic pathways are implicated in microglial-mediated degradation of targeted synapses, we studied CD47 and CD200 expression in the synapse fraction of frozen human cortex. This synapse fraction is highly enriched for the excitatory post-synaptic marker PSD-95, detection of which is absent in the myelin fraction (**Figure 5C**). CD47 abundance was higher in MDD cortical synaptosomes than controls (p=0.0396), whereas CD200 was unchanged (**Figure 5D/E)**.

**Figure 5.**
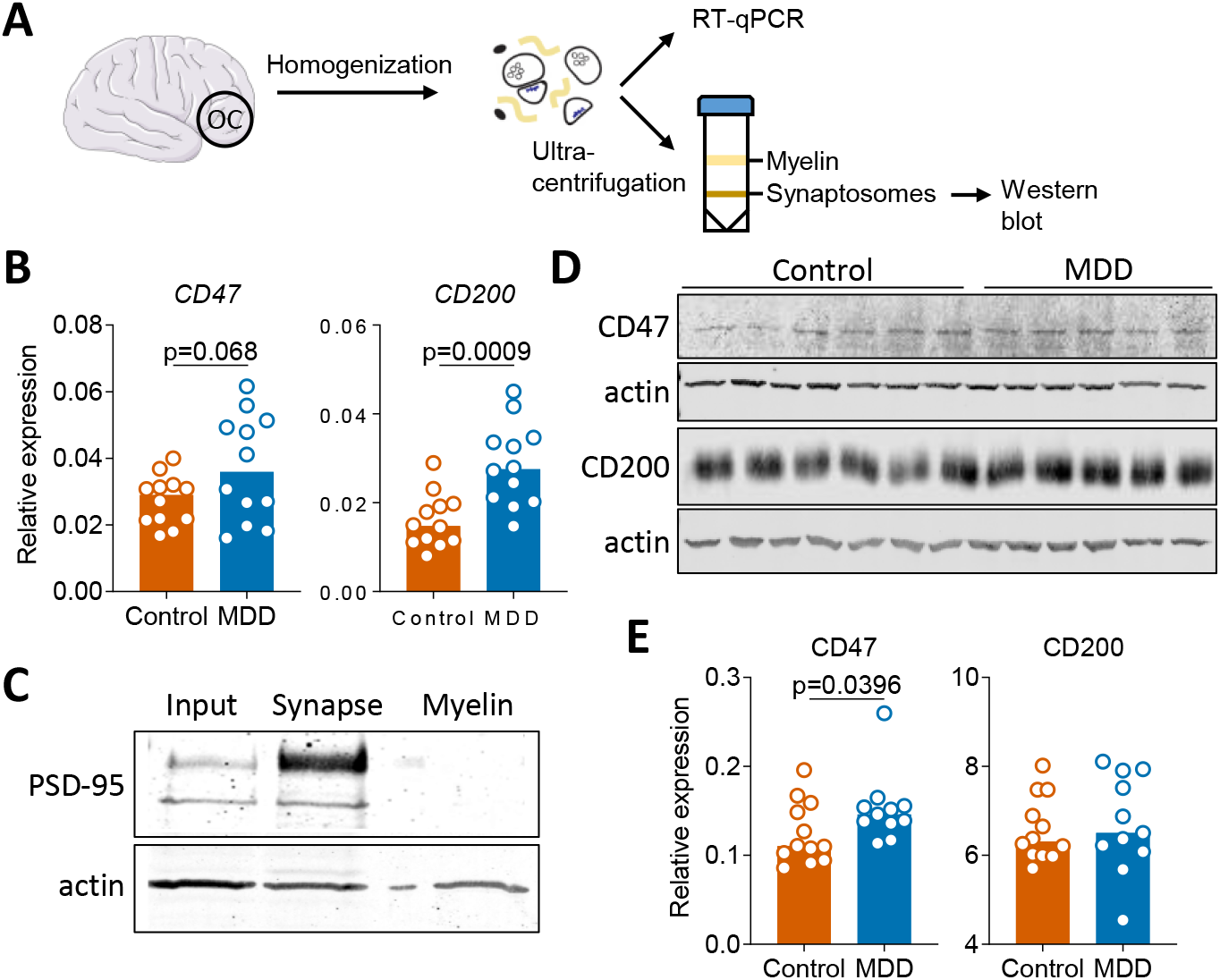
Increased suppression of microglia in MDD cortex. (A) An independent validation cohort of 13 MDD and 13 age-matched control donors was selected. Frozen tissue was extracted from the occipital cortex (OC) for whole tissue RT-qPCR and Western blot analysis of synaptosomes. (B) Expression of CD200 and CD47 in MDD whole tissue determined by RT-qPCR. (C) Western blot analysis of PSD-95 expression in whole tissue homogenate (input), isolated synaptic fractions (synapse), and myelin fractions. (D/E) Western blot analysis of CD47 and CD200 expression in isolated synaptic fractions. Mann-Whitney U test: *p<0.05, ***p<0.001.

## Discussion

Here, we show that GM microglia, but not WM microglia, from brain donors with the clinical diagnosis MDD, show a distinct disease associated transcriptomic profile. The majority of the DE genes (81 out of 92) of GM microglia were downregulated, with numerous genes involved in immune responses (*MK167, SPP1, C1QA/B/C*) and phagocytic function (*FCGR1A/C, FCGR3A, CD14, CD163*). In line with individual genes, pathway analyses showed alterations in immune activation (complement system, iNOS signaling, IL-12 signaling) and phagocytic activity (regulation of phagocytosis, membrane invagination, phagocytosis, FcγR signaling in phagocytosis, IL-12, TLR, TREM1 signaling). An immune-suppressed state of MDD GM microglia is also apparent from a lower CD45 membrane expression compared to control donors, assessed immediately after isolation.

Purified CD11b^+^ cells expressed distinct microglial signature genes but lacked expression of genes defining macrophages, other glial lineages, or neurons(20). The homeostatic signature of GM microglia was unchanged in MDD, based on the unaltered expression of *ADGRG1, CSF1R, P2RY12* and *CX3CR1*. Furthermore, there were no signs of immune activation, such as increased expression of pro-inflammatory cytokines *(IL1, IL6, TNF)*.

The MDD associated microglia (MDD-DAM) with an immune-suppressed phenotype reported here are in line with the results of several recent post-mortem studies of MDD donors(12,13,32–34) and may shed light on the underlying cause of failure of anti-inflammatory treatment trials in MDD(35,36). Snijders et al. (2020) analyzed gene expression in microglia isolated from the medial frontal gyrus, superior temporal gyrus, thalamus, and subventricular zone in MDD using RT-qPCR and also identified a non-inflammatory signature, based on reduced *CD163* and *CD14* expression but enhanced homeostatic gene (*TMEM119* and *CX3CR1*) activity(13). Böttcher et al. (2020) found in a single cell mass cytometry study a non-inflammatory microglia phenotype in MDD characterized by reduced HLA and CD68 expression and an increased homeostatic phenotype(12). Other studies also reported lower expression of microglial inflammatory genes in MDD dorsolateral prefrontal cortex tissue(33) and decreased expression of gene sets involved in endocytosis and antigen processing in the hippocampus and orbitofrontal cortex in MDD(32). As far as we know, our study is the first to examine the transcriptome of acutely isolated post-mortem microglia of MDD cortex, emphasizing the unbiased detection of pathways specifically implicated inMDD-DAM.

Together, these data are in contrast to earlier clinical findings using for example positron emission tomography (PET) imaging with radioligands for the TSPO receptor. This method is frequently applied to indicate neuroinflammation in vivo. TSPO binding was reported to be increased in several brain regions in MDD(19,37,38). However, the sensitivity and specificity of this method for detecting microglial activation in psychiatric disorders have been questioned(39,40). It should be emphasized that our autopsy cohort consists of MDD cases in a late phase, and it could well be that inflammation and microglia activation does occur in earlier stages of the disease.

We only detected transcriptional differences in GM microglia in MDD versus controls and not in subcortical WM. This led us to the hypothesis that changes in microglia might be induced by neuronal interference in MDD. Indeed, we here show increased expression of the neuronal ‘don’t eat me’ signaling molecules CD200 and CD47 in tissue (CD200) and synaptosome fractions (CD47), potentially contributing to the dampened activation status of GM microglia in MDD(41,42). While neuroimmunomodulatory roles of CD47 and CD200 are established, their influence on depressive symptoms has not been studied before. Given that the complement system and phagocytic functions are downregulated in cortical GM microglia in MDD, this may have implications for complement-mediated pruning of synapses. CD47 protects synapses from excess microglia-mediated pruning through signaling via the SIRP1α receptor on microglia during development, and knockout of either CD47 or its receptor results in reduced synapse numbers(29). Also, binding of CD200 to its receptor on myeloid cells (CD200R) is known to control microglial activity(30), and recently, it has been shown that CD200 deficiency leads to synaptic deficits and cognitive dysfunction(43,44). Microglia play an essential role in synaptic circuit remodeling and phagocytosis of synaptic material, and defects may contribute to synaptic abnormalities seen in both neurodevelopmental and neurodegenerative diseases(45,46). Pre-clinical studies in depressed-like rodents showed microglial depletion with attenuated antidepressant and neurogenesis-enhancing treatment effects with imipramine or electroconvulsive stimulation(47). Also, microglial stimulation in chronic unpredictable stress-exposed mice with lipopolysaccharide or macrophage colony-stimulating factor reversed depressed-like behavior(48). One of the most consistent findings in MDD pathology is the deficiency in functional synapses, resulting in disruption of the neural circuits that underlie the regulation of mood(49–51). Chronic stress causes atrophy of neuronal processes and decreases in synaptic density, which is restored by administering newer rapid-acting antidepressants, such as esketamine(52,53).

Concerning limitations, almost all of the MDD brain donors were treated with anti-depressants, and we cannot rule out that these may have influenced the inflammatory status of microglia(54–57). Based on the lack of transcriptomic changes in WM microglia in MDD cases, it seems unlikely that the changes we describe in GM microglia are a direct anti-inflammatory effect of medication. However, a secondary effect of medication through neuronal changes is possible. Controls and MDD brain donors were matched for age, PMD, and pH; however, the age did differ in the RNAseq group (75.0 vs. 63.7, p=0.067), albeit not significant. Furthermore, information on the clinical state of depression at the time of death was unavailable for some donors. This may be important, as it is suggested that synaptic and neuroinflammatory changes in MDD may be state- and severity-dependent(49). However, more than half of our cohort deceased in a depressive state by euthanasia for refractory depression or suicide (7 out of 13; **Supplementary Table 1)**.

In summary, we found indications for an immune-suppressed phenotype of MDD-DAM in the occipital cortex, possibly caused by neuronal regulation. An ‘apathic’ microglial status with reduced phagocytosis and complement activation may have important consequences for synapse metabolism and connectivity relating to MDD. The immune-suppressed MDD-DAM further adds to the continuum of microglia states identified in the human brain.

## Data Availability

All data produced in the present study are available upon reasonable request to the authors

## Acknowledgments

We thank the Psychiatric Donor Program of the Netherlands Brain Bank (NBB-PSY) team for their excellent support and the donors for their valuable donation. A preprint of this manuscript was posted on medRxiv.

## Data availability

After acceptance, the RNA-sequencing dataset is available online in the Gene Expression Omnibus (GEO) database.

---

**Supplementary Table 1.** List of donors used for RNA sequencing of microglia isolated from control or MDD GM or WM.

| Donor | Region | Diagnosis | Sex | Age | PMD | pH CSF | Cause of death | RQN |
| --- | --- | --- | --- | --- | --- | --- | --- | --- |
| 1 | CC, OC | MDD | m | 61-70 | 8:55 | 6.8 | sudden death (pneumonia or cardiac arrest) | 4.8 |
| 2 | CC, OC | control | f | 71-80 | 7:10 | 6.3 | euthanasia for somatic disease (infarction) | 5.5 |
| 3 | CC | MDD | f | 81-90 | 10:00 | 6.3 | burns, dehydration | 4.7 |
| 4 | CC, OC | control | m | 61-70 | 8:45 | 6.4 | pharyngeal cancers | 6.2 |
| 5 | CC, OC | MDD | m | 41-50 | 5:55 | 6.7 | euthanasia for psychiatric disorder | 5.8 |
| 6 | CC, OC | control | m | 61-70 | 5:50 | 6.5 | cancer | 5.1 |
| 7 | CC, OC | MDD | m | 51-60 | 4:30 | 6.8 | euthanasia for psychiatric disorder | 7.8 |
| 8 | CC, OC | control | m | 81-90 | 8:12 | 6.6 | heart failure | 6.6 |
| 9 | CC, OC | control | f | 81-90 | 7:40 | 7.5 | sepsis, terminal renal insufficiency | 5.0 |
| 10 | CC, OC | control | m | 81-90 | 5:05 | 7.1 | euthanasia for somatic disease (urothelial carcinoma) | 5.5 |
| 11 | CC, OC | MDD | f | 61-70 | 6:25 | 6.3 | euthanasia for somatic disease (colon carcinoma) | 4.5 |
| 12 | CC, OC | MDD | f | 61-70 | 6:55 | 6.3 | congestive heart failure | 4.5 |
| 13 | CC, OC | MDD | f | 71-80 | 8:40 | 6.8 | euthanasia for psychiatric disorder | 5.9 |
| 14 | CC, OC | MDD | m | 41-50 | 9:40 | 6.8 | euthanasia for psychiatric disorder | 7.1 |
| 15 | CC, OC | control | f | 51-60 | 5:30 | 7.1 | euthanasia for somatic disease (mamma carcinoma) | 6.8 |
| 16 | CC, OC | MDD | f | 61-70 | 8:40 | 7.1 | euthanasia for psychiatric disorder | 4.3 |
| 17 | CC, OC | MDD | m | 41-50 | 12:45 | 6.8 | euthanasia for psychiatric disorder | 4.2 |
| 18 | OC | control | m | 71-80 | 9:30 | n/a | respiratory failure due to traumatic cervical paraplegia | 3.9 |
| 19 | CC, OC | MDD | f | 81-90 | 4:30 | 6.9 | euthanasia for somatic disease (thalamus infarction) | 6.2 |
| 20 | CC, OC | MDD | m | 61-70 | 5:45 | n/a | euthanasia for somatic disease (multiple infarctions) | 6.2 |
| 21 | CC, OC | control | f | 81-90 | 6:50 | 6.8 | euthanasia for somatic disease (multi morbidity) | 6.2 |
| 22 | CC, OC | control | f | 51-60 | 7:30 | n/a | euthanasia for somatic disease (severe pain) | 7.3 |
| 23 | OC | MDD | f | 41-50 | 11:20 | n/a | suicide | 5.5 |

Age: age at death (years), PMD: post-mortem delay (hours), CSF: cerebrospinal fluid, CC: corpus callosum (WM), OC: occipital cortex (GM), MDD: major depressive disorder, f: female, m: male, n/a: data not available, RQN: RNA quality number.

**Supplementary Table 2.**
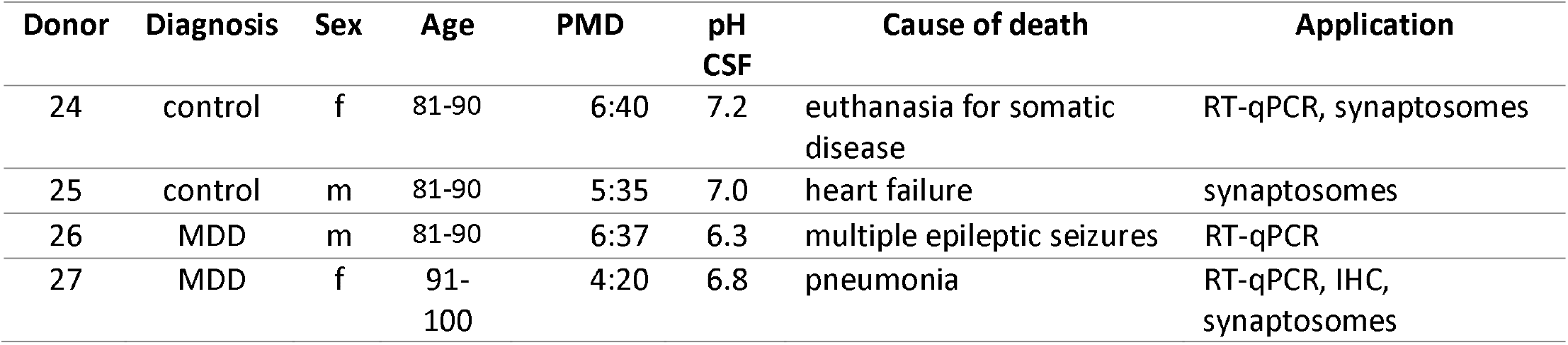
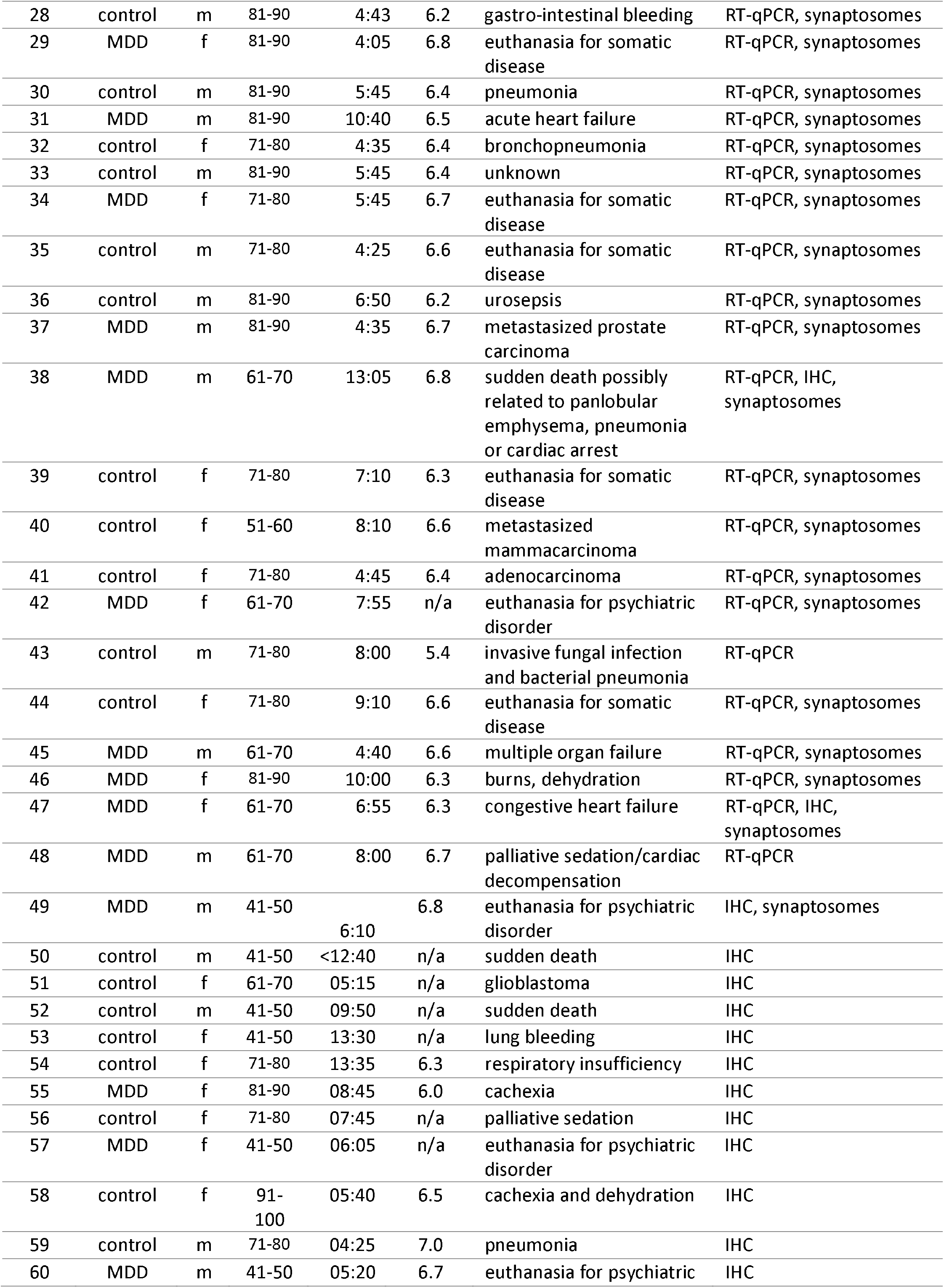
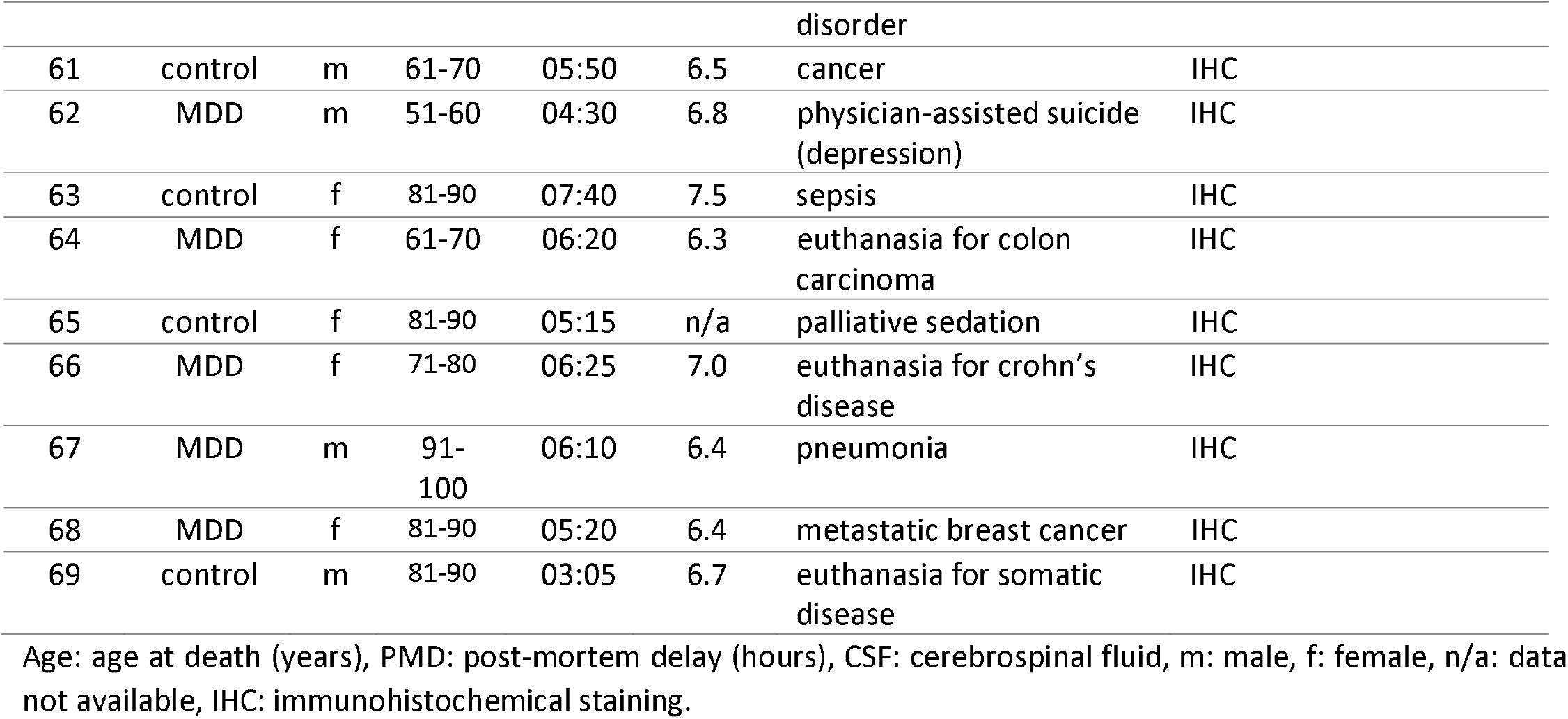
List of donors used for additional characterization of microglia from control or MDD GM; RT-qPCR, IHC or isolation of synaptosomes from fresh-frozen GM tissue.

**Supplementary Table 3.**
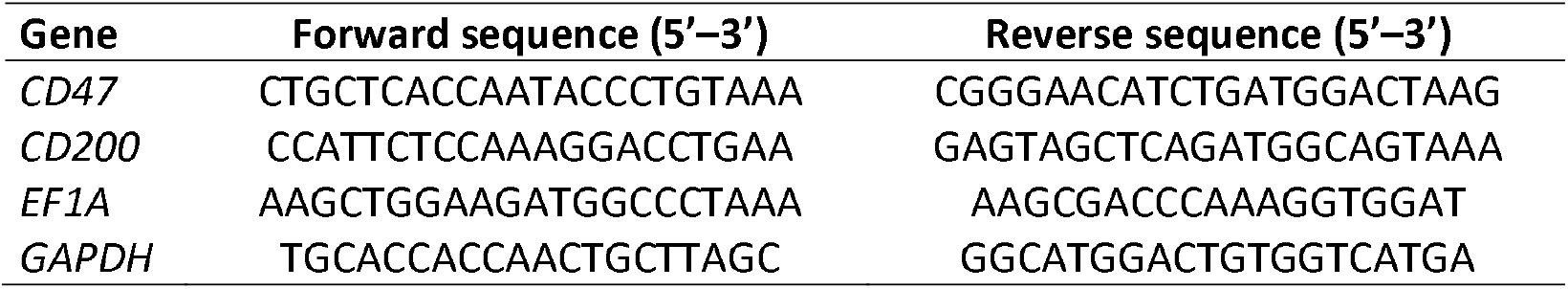
List of primers used for RT-qPCR.

**Supplementary Table 4.**
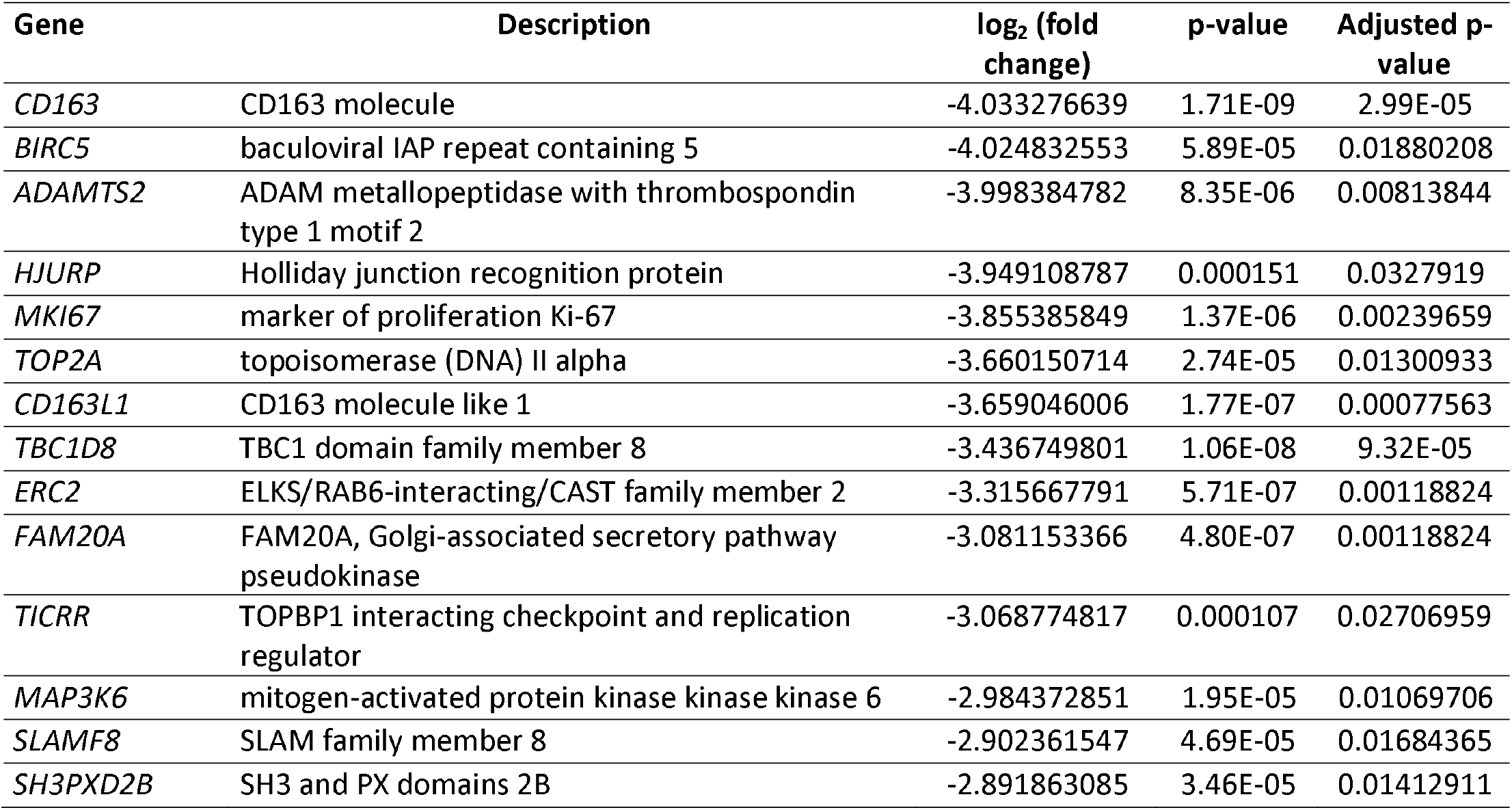
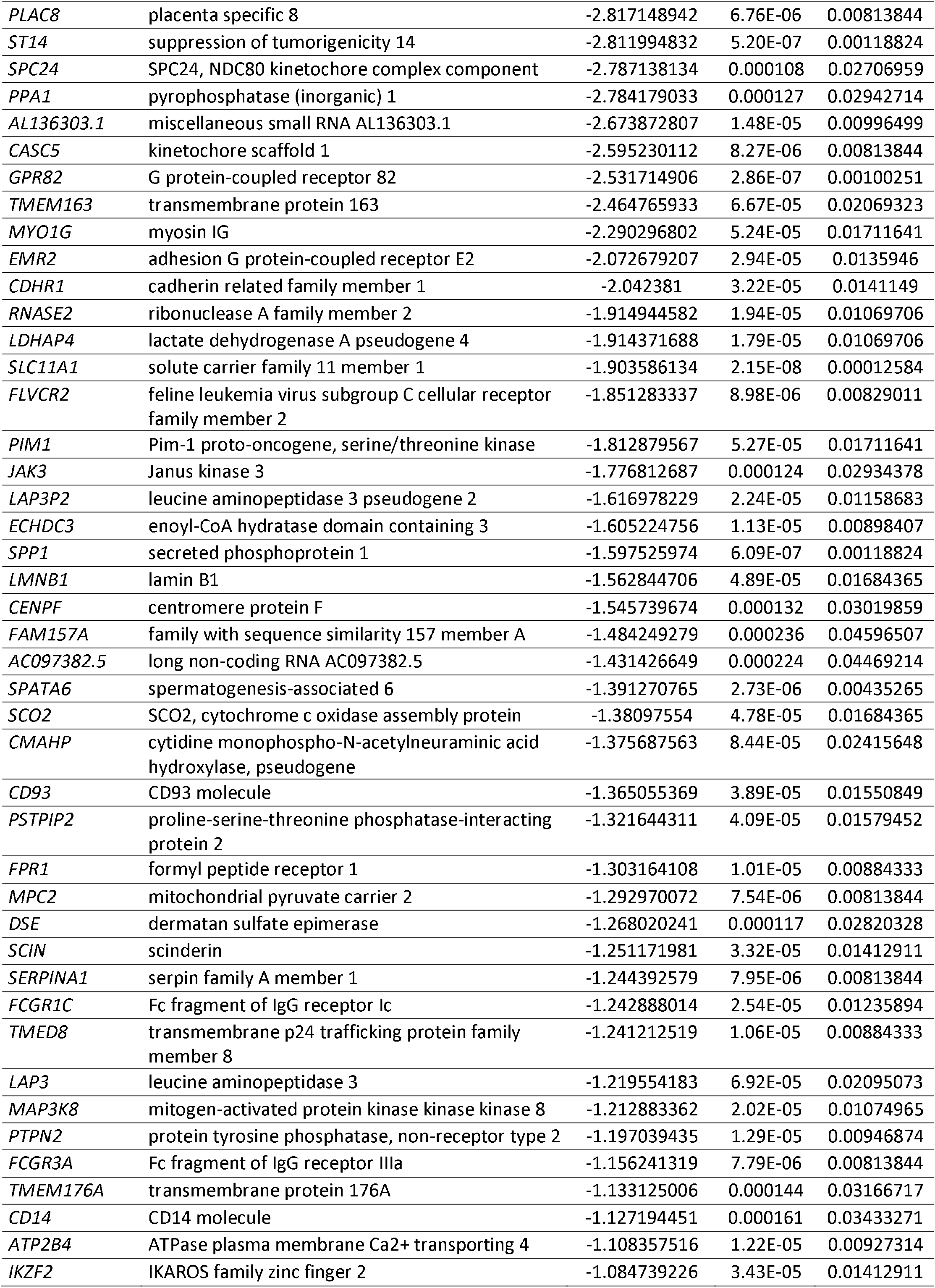
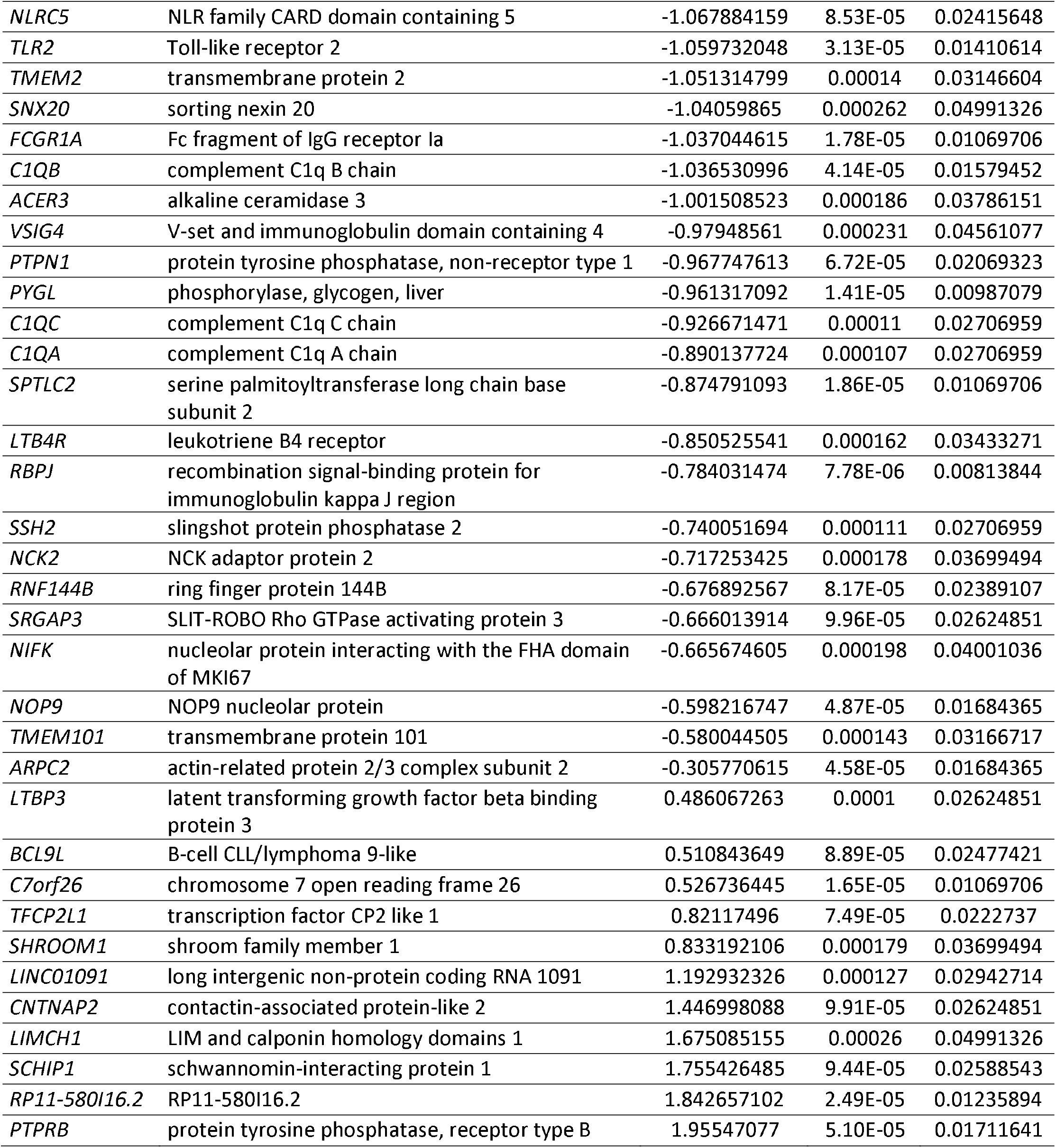
Complete list of DE genes as determined by RNA sequencing of microglia isolated from control or MDD GM. Negative log2 (fold change) values indicate downregulation of a gene in MDD microglia compared to control microglia.

**Supplementary Table 5.** Gene score resampling (GSR) of MDD cortical microglia gene expression compared to control microglia.

| GO term | Description | Size | Multi F. | Corrected p-value |
| --- | --- | --- | --- | --- |
| GO:0019864 | IgG binding | 9 | 0.0302 | 8.14E-09 |
| GO:0042116 | macrophage activation | 32 | 0.855 | 1.46E-08 |
| GO:0019724 | B cell-mediated immunity | 63 | 0.859 | 0.00000266 |
| GO:0038094 | Fc-gamma receptor signaling pathway | 72 | 0.843 | 0.00000262 |
| GO:0016064 | immunoglobulin-mediated immune response | 62 | 0.861 | 0.00000532 |
| GO:0101003 | Ficolin 1-rich granule membrane | 59 | 0.64 | 0.00000491 |
| GO:0002455 | humoral immune response mediated by circulating immunoglobulin | 32 | 0.655 | 0.0000213 |
| GO:0006953 | acute-phase response | 32 | 0.696 | 0.0000403 |
| GO:0038096 | Fc-gamma receptor signaling pathway involved in phagocytosis | 68 | 0.843 | 0.0000446 |
| GO:0050663 | cytokine secretion | 39 | 0.852 | 0.0000953 |
| GO:0050764 | regulation of phagocytosis | 71 | 0.885 | 0.0000972 |
| GO:0099024 | plasma membrane invagination | 43 | 0.764 | 0.000137 |
| GO:0033116 | endoplasmic reticulum-Golgi intermediate compartment membrane | 66 | 0.449 | 0.000266 |
| GO:0010324 | membrane invagination | 50 | 0.786 | 0.000335 |
| GO:0046915 | transition metal ion transmembrane transporter activity | 38 | 0.618 | 0.000536 |
| GO:0030670 | phagocytic vesicle membrane | 68 | 0.87 | 0.000522 |
| GO:0006911 | phagocytosis, engulfment | 34 | 0.703 | 0.000558 |
| GO:0002102 | podosome | 34 | 0.533 | 0.000558 |
| GO:0006879 | cellular iron ion homeostasis | 60 | 0.838 | 0.0016 |
| GO:0030449 | regulation of complement activation | 41 | 0.71 | 0.00192 |

GSR was performed using ErmineJ. GO was set between 5 and 75 genes, 200,000 iterations, false discovery rate (FDR) <0.001.

**Supplementary Figure 1.**
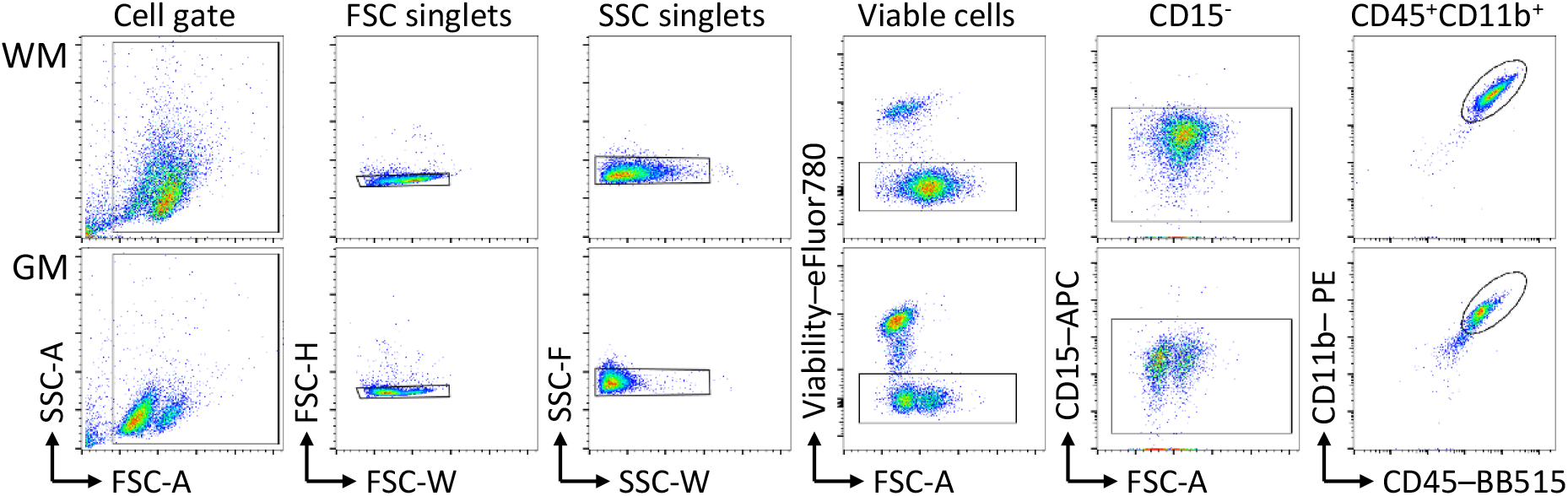
Flow cytometry gating strategy for human microglia in white matter (WM) and gray matter (GM). Representative dot plots showing the gating strategy used for Figure 1A with from left to right: cell gating by forward scatter (FCS) and sideward scatter (SSC), FCS-width/height duplet exclusion, SSC-width/height duplet exclusion, gating of viable cells, gating of CD15^-^ events, and gating of CD45^+^CD11b^+^ events.

**Supplementary Figure 1.**
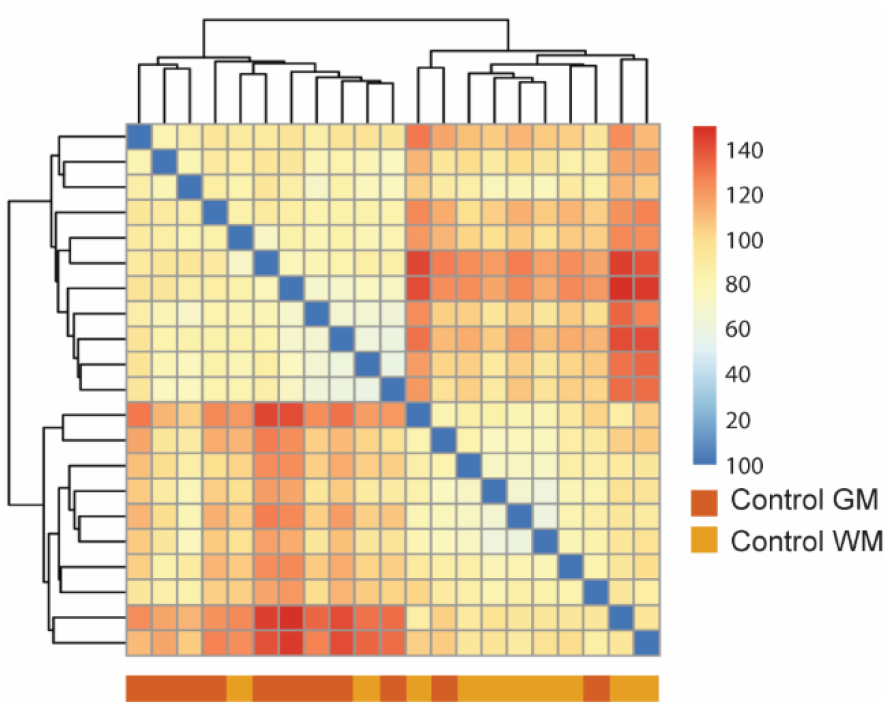
Hierarchical clustering of gene expression data obtained from WM and GM microglia of control brains.

## References

1. Abbafati C, Abbas KM, Abbasi-Kangevari M, Abd-Allah F, Abdelalim A, Abdollahi M, et al. (2020): Global burden of 369 diseases and injuries in 204 countries and territories, 1990–2019: a systematic analysis for the Global Burden of Disease Study 2019. Lancet. https://doi.org/10.1016/S0140-6736(20)30925-9

2. Rush AJ, Trivedi MH, Wisniewski SR, Nierenberg AA, Stewart JW, Warden D, et al. (2006): Acute and longer-term outcomes in depressed outpatients requiring one or several treatment steps: A STAR*D report. Am J Psychiatry. https://doi.org/10.1176/ajp.2006.163.11.1905

3. Köhler CA, Freitas TH, Maes M, de Andrade NQ, Liu CS, Fernandes BS, et al. (2017): Peripheral cytokine and chemokine alterations in depression: a meta-analysis of 82 studies. Acta Psychiatr Scand. https://doi.org/10.1111/acps.12698

4. Mariani N, Cattane N, Pariante C, Cattaneo A (2021): Gene expression studies in Depression development and treatment: an overview of the underlying molecular mechanisms and biological processes to identify biomarkers. Transl Psychiatry 11. https://doi.org/11:354 https://doi.org/10.1038/s41398-021-01469-6

5. Eyre HA, Air T, Pradhan A, Johnston J, Lavretsky H, Stuart MJ, Baune BT (2016): A meta-analysis of chemokines in major depression. Prog Neuro-Psychopharmacology Biol Psychiatry. https://doi.org/10.1016/j.pnpbp.2016.02.006

6. Euesden J, Danese A, Lewis CM, Maughan B (2017): A bidirectional relationship between depression and the autoimmune disorders - New perspectives from the National Child Development Study. PLoS One. https://doi.org/10.1371/journal.pone.0173015

7. Andersson NW, Gustafsson LN, Okkels N, Taha F, Cole SW, Munk-Jorgensen P, Goodwin RD (2015): Depression and the risk of autoimmune disease: A nationally representative, prospective longitudinal study. Psychol Med. https://doi.org/10.1017/S0033291715001488

8. Hoogland ICM, Houbolt C, van Westerloo DJ, van Gool WA, van de Beek D (2015): Systemic inflammation and microglial activation: Systematic review of animal experiments. Journal of Neuroinflammation. https://doi.org/10.1186/s12974-015-0332-6

9. Hoogland ICM, Westhoff D, Engelen-Lee JY, Melief J, Valls Serón M, Houben-Weerts JHMP, et al. (2018): Microglial activation after systemic stimulation with lipopolysaccharide and Escherichia coli. Front Cell Neurosci. https://doi.org/10.3389/fncel.2018.00110

10. Hendrickx DAE, van Scheppingen J, van der Poel M, Bossers K, Schuurman KG, van Eden CG, et al. (2017): Gene expression profiling of multiple sclerosis pathology identifies early patterns of demyelination surrounding chronic active lesions. Front Immunol. https://doi.org/10.3389/fimmu.2017.01810

11. Ramaglia V, Dubey M, Malpede MA, Petersen N, de Vries SI, Ahmed SM, et al. (2021): Complement-associated loss of CA2 inhibitory synapses in the demyelinated hippocampus impairs memory. Acta Neuropathol. https://doi.org/10.1007/s00401-021-02338-8

12. Böttcher C, Fernández-Zapata C, Snijders GJL, Schlickeiser S, Sneeboer MAM, Kunkel D, et al. (2020): Single-cell mass cytometry of microglia in major depressive disorder reveals a non-inflammatory phenotype with increased homeostatic marker expression. Transl Psychiatry 10. https://doi.org/10.1038/s41398-020-00992-2

13. Snijders GJLJ, Sneeboer MAM, Fernández-Andreu A, Udine E, Boks MP, Ormel PR, et al. (2021): Distinct non-inflammatory signature of microglia in post-mortem brain tissue of patients with major depressive disorder. Mol Psychiatry. https://doi.org/10.1038/s41380-020-00896-z

14. Hammond BP, Manek R, Kerr BJ, Macauley MS, Plemel JR (2021): Regulation of microglia population dynamics throughout development, health, and disease. GLIA. https://doi.org/10.1002/glia.24047

15. Sneeboer MAM, Snijders GJLJ, Berdowski WM, Fernández-Andreu A, van Mierlo HC, Berdenis van Berlekom A, et al. (2019): Microglia in post-mortem brain tissue of patients with bipolar disorder are not immune activated. Transl Psychiatry. https://doi.org/10.1038/s41398-019-0490-x

16. Snijders GJLJ, van Zuiden W, Sneeboer MAM, Berdenis van Berlekom A, van der Geest AT, Schnieder T, et al. (2021): A loss of mature microglial markers without immune activation in schizophrenia. Glia. https://doi.org/10.1002/glia.23962

17. Torres-Platas SG, Cruceanu C, Chen GG, Turecki G, Mechawar N (2014): Evidence for increased microglial priming and macrophage recruitment in the dorsal anterior cingulate white matter of depressed suicides. Brain Behav Immun 42: 50–59.

18. Hannestad J, DellaGioia N, Gallezot JD, Lim K, Nabulsi N, Esterlis I, et al. (2013): The neuroinflammation marker translocator protein is not elevated in individuals with mild-to-moderate depression: A [11C]PBR28 PET study. Brain Behav Immun. https://doi.org/10.1016/j.bbi.2013.06.010

19. Holmes SE, Hinz R, Conen S, Gregory CJ, Matthews JC, Anton-Rodriguez JM, et al. (2018): Elevated Translocator Protein in Anterior Cingulate in Major Depression and a Role for Inflammation in Suicidal Thinking: A Positron Emission Tomography Study. Biol Psychiatry. https://doi.org/10.1016/j.biopsych.2017.08.005

20. Mizee MR, Miedema SSM, van der Poel M Adelia, Schuurman KG, van Strien ME, et al. (2017): Isolation of primary microglia from the human post-mortem brain: effects of ante- and postmortem variables. Acta Neuropathol Commun. https://doi.org/10.1186/s40478-017-0418-8

21. Melief J, Koning N, Schuurman KG, Van De Garde MDB, Smolders J, Hoek RM, et al. (2012): Phenotyping primary human microglia: tight regulation of LPS responsiveness. Glia 60: 1506–17.

22. Leek JT, Johnson WE, Parker HS, Jaffe AE, Storey JD (2012): The SVA package for removing batch effects and other unwanted variation in high-throughput experiments. Bioinformatics. https://doi.org/10.1093/bioinformatics/bts034

23. Leek JT (2011): Asymptotic Conditional Singular Value Decomposition for High-Dimensional Genomic Data. Biometrics. https://doi.org/10.1111/j.1541-0420.2010.01455.x

24. Zhou Y, Zhou B, Pache L, Chang M, Khodabakhshi AH, Tanaseichuk O, et al. (2019): Metascape provides a biologist-oriented resource for the analysis of systems-level datasets. Nat Commun. https://doi.org/10.1038/s41467-019-09234-6

25. Carlin RK, Grab DJ, Cohen RS, Siekevitz P (1980): Isolation and characterization of postsynaptic densities from various brain regions: Enrichment of different types of postsynaptic densities. J Cell Biol. https://doi.org/10.1083/jcb.86.3.831

26. van der Poel M, Ulas T, Mizee MR, Hsiao CC, Miedema SSM Adelia, et al. (2019): Transcriptional profiling of human microglia reveals grey–white matter heterogeneity and multiple sclerosis-associated changes. Nat Commun. https://doi.org/10.1038/s41467-019-08976-7

27. Masuda T, Sankowski R, Staszewski O, Böttcher C, Amann L Sagar, et al. (2019): Spatial and temporal heterogeneity of mouse and human microglia at single-cell resolution. Nature. https://doi.org/10.1038/s41586-019-0924-x

28. Absinta M, Maric D, Gharagozloo M, Garton T, Smith MD, Jin J, et al. (2021): A lymphocyte– microglia–astrocyte axis in chronic active multiple sclerosis. Nature. https://doi.org/10.1038/s41586-021-03892-7

29. Lehrman EK, Wilton DK, Litvina EY, Welsh CA, Chang ST, Frouin A, et al. (2018): CD47 Protects Synapses from Excess Microglia-Mediated Pruning during Development. Neuron 100: 120-134.e6.

30. Barclay AN, Wright GJ, Brooke G, Brown MH (2002, June): CD200 and membrane protein interactions in the control of myeloid cells. Trends in Immunology, vol. 23. Trends Immunol, pp 285–290.

31. Elward K, Gasque P (2003): “Eat me” and “don’t eat me” signals govern the innate immune response and tissue repair in the CNS: Emphasis on the critical role of the complement system. Molecular Immunology, vol. 40 40: 85–94.

32. Darby MM, Yolken RH, Sabunciyan S (2016): Consistently altered expression of gene sets in postmortem brains of individuals with major psychiatric disorders. Transl Psychiatry 6: e890.

33. Pantazatos SP, Huang Y-Y, Rosoklija GB, Dwork AJ, Arango V, Mann JJ (2017): Whole-transcriptome brain expression and exon-usage profiling in major depression and suicide: evidence for altered glial, endothelial and ATPase activity. Mol Psychiatry 22: 760–773.

34. Paolicelli RC, Sierra A, Stevens B, Tremblay ME, Aguzzi A, Ajami B, et al. (2022): Microglia states and nomenclature: A field at its crossroads. Neuron 110: 3458–3483.

35. Miller AH, Pariante CM (2020): Trial failures of anti-inflammatory drugs in depression. The Lancet Psychiatry. https://doi.org/10.1016/S2215-0366(20)30357-6

36. Raison CL, Rutherford RE, Woolwine BJ, Shuo C, Schettler P, Drake DF, et al. (2013): A randomized controlled trial of the tumor necrosis factor antagonist infliximab for treatment-resistant depression: The role of baseline inflammatory biomarkers. Arch Gen Psychiatry. https://doi.org/10.1001/2013.jamapsychiatry.4

37. Li H, Sagar AP, Kéri S (2018): Translocator protein (18 kDa TSPO) binding, a marker of microglia, is reduced in major depression during cognitive-behavioral therapy. Prog Neuro-Psychopharmacology Biol Psychiatry 83: 1–7.

38. Setiawan E, Attwells S, Wilson AA, Mizrahi R, Rusjan PM, Miler L, et al. (2018): Association of translocator protein total distribution volume with duration of untreated major depressive disorder: a cross-sectional study. The Lancet Psychiatry 5: 339–347.

39. Sneeboer MAM, van der Doef T, Litjens M, Psy NBB, Melief J, Hol EM, et al. (2020): Microglial activation in schizophrenia: Is translocator 18l1kDa protein (TSPO) the right marker? Schizophr Res 215: 167–172.

40. Owen DR, Narayan N, Wells L, Healy L, Smyth E, Rabiner EA, et al. (2017): Pro-inflammatory activation of primary microglia and macrophages increases 18 kDa translocator protein expression in rodents but not humans. J Cereb Blood Flow Metab 37: 2679–2690.

41. Biber K, Neumann H, Inoue K, Boddeke HWGM (2007): Neuronal “On” and “Off” signals control microglia. Trends in Neurosciences. https://doi.org/10.1016/j.tins.2007.08.007

42. Koning N, Swaab DF, Hoek RM, Huitinga I (2009): Distribution of the immune inhibitory molecules CD200 and CD200R in the normal central nervous system and multiple sclerosis lesions suggests neuron-glia and glia-glia interactions. J Neuropathol Exp Neurol. https://doi.org/10.1097/NEN.0b013e3181964113

43. Ting AY, Stawski PS, Draycott AS, Udeshi ND, Lehrman EK, Wilton DK, et al. (2016): Proteomic Analysis of Unbounded Cellular Compartments: Synaptic Clefts. Cell 166: 1295-1307.e21.

44. Feng D, Huang A, Yan W, Chen D (2019): CD200 dysfunction in neuron contributes to synaptic deficits and cognitive impairment. Biochem Biophys Res Commun 516: 1053–1059.

45. Paolicelli RC, Bolasco G, Pagani F, Maggi L, Scianni M, Panzanelli P, et al. (2011): Synaptic pruning by microglia is necessary for normal brain development. Science (80-) 333: 1456–1458.

46. Schafer DP, Lehrman EK, Kautzman AG, Koyama R, Mardinly AR, Yamasaki R, et al. (2012): Microglia Sculpt Postnatal Neural Circuits in an Activity and Complement-Dependent Manner. Neuron 74: 691–705.

47. Rimmerman N, Verdiger H, Goldenberg H, Naggan L, Robinson E, Kozela E, et al. (2022): Microglia and their LAG3 checkpoint underlie the antidepressant and neurogenesis-enhancing effects of electroconvulsive stimulation. Mol Psychiatry. https://doi.org/10.1038/s41380-021-01338-0

48. Kreisel T, Frank MG, Licht T, Reshef R, Ben-Menachem-Zidon O, Baratta M V., et al. (2014): Dynamic microglial alterations underlie stress-induced depressive-like behavior and suppressed neurogenesis. Mol Psychiatry. https://doi.org/10.1038/mp.2013.155

49. Holmes SE, Scheinost D, Finnema SJ, Naganawa M, Davis MT, DellaGioia N, et al. (2019): Lower synaptic density is associated with depression severity and network alterations. Nat Commun. https://doi.org/10.1038/s41467-019-09562-7

50. Duman RS, Aghajanian GK, Sanacora G, Krystal JH (2016): Synaptic plasticity and depression: New insights from stress and rapid-acting antidepressants. Nature Medicine. https://doi.org/10.1038/nm.4050

51. Kang HJ, Voleti B, Hajszan T, Rajkowska G, Stockmeier CA, Licznerski P, et al. (2012): Decreased expression of synapse-related genes and loss of synapses in major depressive disorder. Nat Med. https://doi.org/10.1038/nm.2886

52. Li N, Lee B, Liu RJ, Banasr M, Dwyer JM, Iwata M, et al. (2010): mTOR-dependent synapse formation underlies the rapid antidepressant effects of NMDA antagonists. Science (80-). https://doi.org/10.1126/science.1190287

53. Liu RJ, Lee FS, Li XY, Bambico F, Duman RS, Aghajanian GK (2012): Brain-derived neurotrophic factor Val66Met allele impairs basal and ketamine-stimulated synaptogenesis in prefrontal cortex. Biol Psychiatry. https://doi.org/10.1016/j.biopsych.2011.09.030

54. Basterzi AD, Aydemir Ç, Kisa C, Aksaray S, Tuzer V, Yazici K, Göka E (2005): IL-6 levels decrease with SSRI treatment in patients with major depression. Hum Psychopharmacol. https://doi.org/10.1002/hup.717

55. Ghareghani M, Zibara K, Sadeghi H, Dokoohaki S, Sadeghi H, Aryanpour R, Ghanbari A (2017): Fluvoxamine stimulates oligodendrogenesis of cultured neural stem cells and attenuates inflammation and demyelination in an animal model of multiple sclerosis. Sci Rep. https://doi.org/10.1038/s41598-017-04968-z

56. Tynan RJ, Weidenhofer J, Hinwood M, Cairns MJ, Day TA, Walker FR (2012): A comparative examination of the anti-inflammatory effects of SSRI and SNRI antidepressants on LPS stimulated microglia. Brain Behav Immun. https://doi.org/10.1016/j.bbi.2011.12.011

57. Faissner S, Mishra M, Kaushik DK, Wang J, Fan Y, Silva C, et al. (2017): Systematic screening of generic drugs for progressive multiple sclerosis identifies clomipramine as a promising therapeutic. Nat Commun. https://doi.org/10.1038/s41467-017-02119-6

